# Text Mining Approach to Analyze Coronavirus Impact: Mexico City as Case of Study

**DOI:** 10.1101/2020.05.07.20094466

**Authors:** Josimar Edinson Chire Saire, Anabel Pineda Briseño

## Abstract

The epidemiological outbreak of a novel coronavirus (2019-nCoV or Covid-19) in China, and its rapid spread, gave rise to the first pandemic in the digital age. Derived from this fact that has surprised humanity, many countries started with different strategies in order to stop the infection. In this context, one of the greatest challenges for the scientific community is monitoring (real time) the global population to get immediate feedback of what is happening with the people during this public health contingency. An alternative interesting and affordable for the materialization of the aforementioned are the social networks. In a social network, the persons can act as sensors/information not only of personal data but also data derived from their behavior. This paper aims to analyze the publications of people in Mexico using a Text Mining approach. Specifically, Mexico City is presented as a case study to help understand the impact on society of the spread of Covid-19.

## I. Introduction

A novel infectious disease (2019-nCoV or better known as Covid-19) identified at the end of 2019 in Wuhan, China, originated an outbreak of viral pneumonia earlier this year. Due the rapid dissemination of this respiratory disease, increased deaths and resource depletion, the World Health Organization (WHO) declared coronavirus as pandemic [1] [2]. This international public health emergency is rapidly mobilizing the government, the industry and the scientific community to respond effectively to this new disease that represents a highrisk for human population and a negative impact for others areas such as economy, education, politics, etc. By April 30, 2020, 3,096,626 confirmed cases of Covid-19 and 217,896 deaths were globally reported to World Health Organization (WHO) [3]. On February 29, 2020, the first two cases of Covid-19 were reported in Mexico. By April 30, the total number of infections was 17,799 and 1732 deaths in this North American country [4]. Regarding Mexico City, by April 30, 2020, there have been confirmed 6412 cases and around of 379 persons have died due to the Covid-19 disease [5].

Over the past few years, the explosive growth of Social Networks (SNs) indicates that more people are using them to connect and communicate their ideas, interests, feelings, experiences, events, including health related information. Therefore, the SNs are a huge source of heterogeneous data can be exploited for public health monitoring and surveillance purposes. [6] [7]. Among SNs, ”Twitter” is one of the most widely used micro-blogs. It’s a free service for sharing short-text messages or ”tweets” limited to 280 characters. Currently in the world Twitter has around 340 million active users [8]. Meanwhile in Mexico, with a population of around 125 million people, it has an audience of 9.45 million of users, being Mexico City with the highest number of active twitter users [9].

The main contribution of this work is evaluate the keywords related to Covid-19 in an effort to understand how a public health emergency of international concern plays out in social media, and Twitter in particular, in Mexico City.

The remainder of the paper follows. Section 2 presents related works regarding the retrieval infectious diseases information from social media. In section 3, the data collection methodology for extracting relevant information of Covid-19 from Twitter is presented. Section 4 describes experimental findings and a discussion related to the analysis. Finally, conclusions and future work are described in Section 5.

## II. Related work

There are many research papers discussing the uses of Twitter data, and the valuable contribution of information for the field of public health. In this section some important approaches are summarized. In the works [10] and [11] are presented studies about the power of using Natural Language Processing (NLP) techniques to generate new information collected from Twitter for public health research. other contributions, such as [12], demonstrated experimental findings that give rise to future research works on when to use Twitter information for public health. In order to show the effectiveness of use social media information as strategy for public research in [13] and [7] are presented some analysis. Both research papers conclude with a recommendation to combine social media information with other techniques to measure disease surveillance and spread. In [14] is presented a real time system for the prediction and detection of the proliferation of an epidemic by identifying disease tweets by graphical location. A seminal paper published by [15] proposes combining Twitter data with Goolgle Trends data to track the spread of infectious diseases. A study conducted by [16] analyzed Twitter data collected during some infectious disease outbreaks. The experimental findings helped to learn more about how act people with panic disorder into a health contingency. Other interesting proposals are related to applications for surveillance with data source from Twitter. For example: monitor H1N1 pandemic [17], monitor Dengue in Brazil [18], monitor Covid-19 symptoms in Bogota, Colombia [19], and monitor Covid-19 in South American Population [20].

The previous works have examined Twitter content around infectious diseases, such as Ebola, Dengue, N1H1, Covid-19, etc. Because there is very little scientific literature that has conducted a analysis of how the dissemination of the new coronavirus Covid-19 is affecting in different aspects to the society, in this paper a text mining approach is proposed to analyze the impact of Covid-19 in the Mexican society, particularly in the population of Mexico City, one of the most populated cities in the world and in Latin America.

## III. Methodology

The present work performs experiments with source data from Twitter with Natural Language Processing and Data Mining (Text Mining).

- Choose terms to search on Twitter
- Setup parameters of the query for Twitter and collect data
- Pre-processing data to eliminate words with no relevance (stopwords)
- Visualization

### A. Gather Relevant Terms

Considering news about Covid-19 and after previous queries.The next terms are chosen:

- ’coronavirus’,’covid19’

But, people does not write following this official names then special characters are found like @, #, -, _. For this reason, variations of the previous terms are created, i.e. { ’@coronavirus’, #covid-19’, ’@covid_19’ }

### B. Setup Parameters for the Query and Collect Data

The extraction of tweets is through Twitter API, with the next parameters:

- date: 13-03-2020 to 20-03-2020
- terms: the chosen words mentioned in previous subsection
- geolocalization: Mexico City (Capital of Mexico). Fig. 1 7 (19.4333,-99.1333)
- continent: North America
- language: Spanish
- radius: around 50 km

**Fig. 1.**
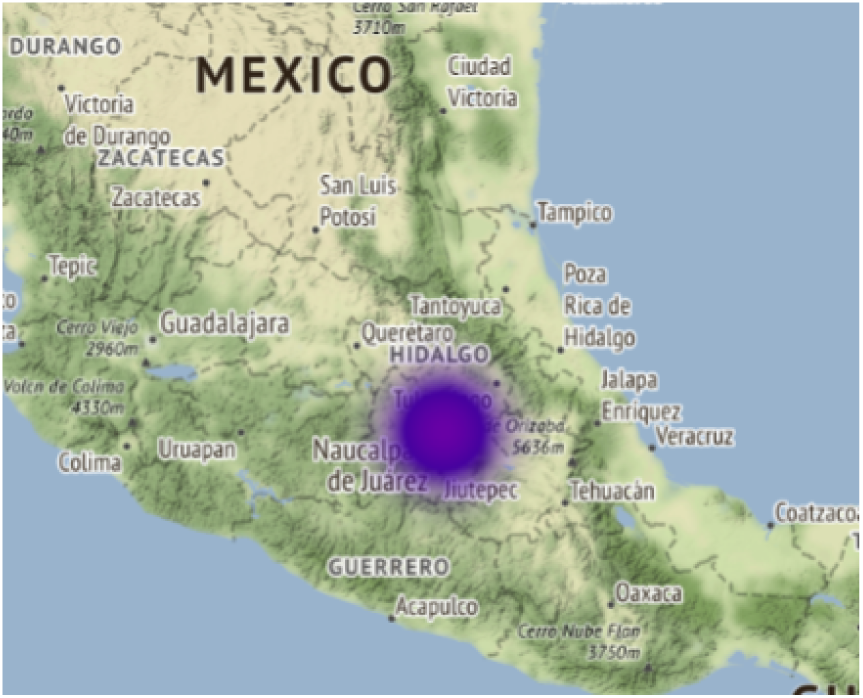
Geolocalization of Mexico City

### C. Preprocessing Data

- Uppercase to lowercase
- Eliminate alphanumeric symbols
- Eliminate words with size less or equal than 3
- Add some exceptions

### D. Visualization

- The number of infected people per day and accumulated to show the progress
- Tweets per day to analyze the increasing number of posts
- Cloud of words to analyze the most frequent terms involved both per day and per a time period

## IV. Results

The next graphics present the results of the experiments and answer some questions to understand the phenomenon of the pandemic over Mexico City population.

### A. How was the progress of Covid-19 in Mexico City?

The Figure 2 shows the progress of positive cases per day and accumulated of Covid-19 in Mexico City from the first outbreaks that were reported in late February to April 30, 2020 [5]. As can be seen, positive cases began to increase steadily from the third week of March.

**Fig. 2.**
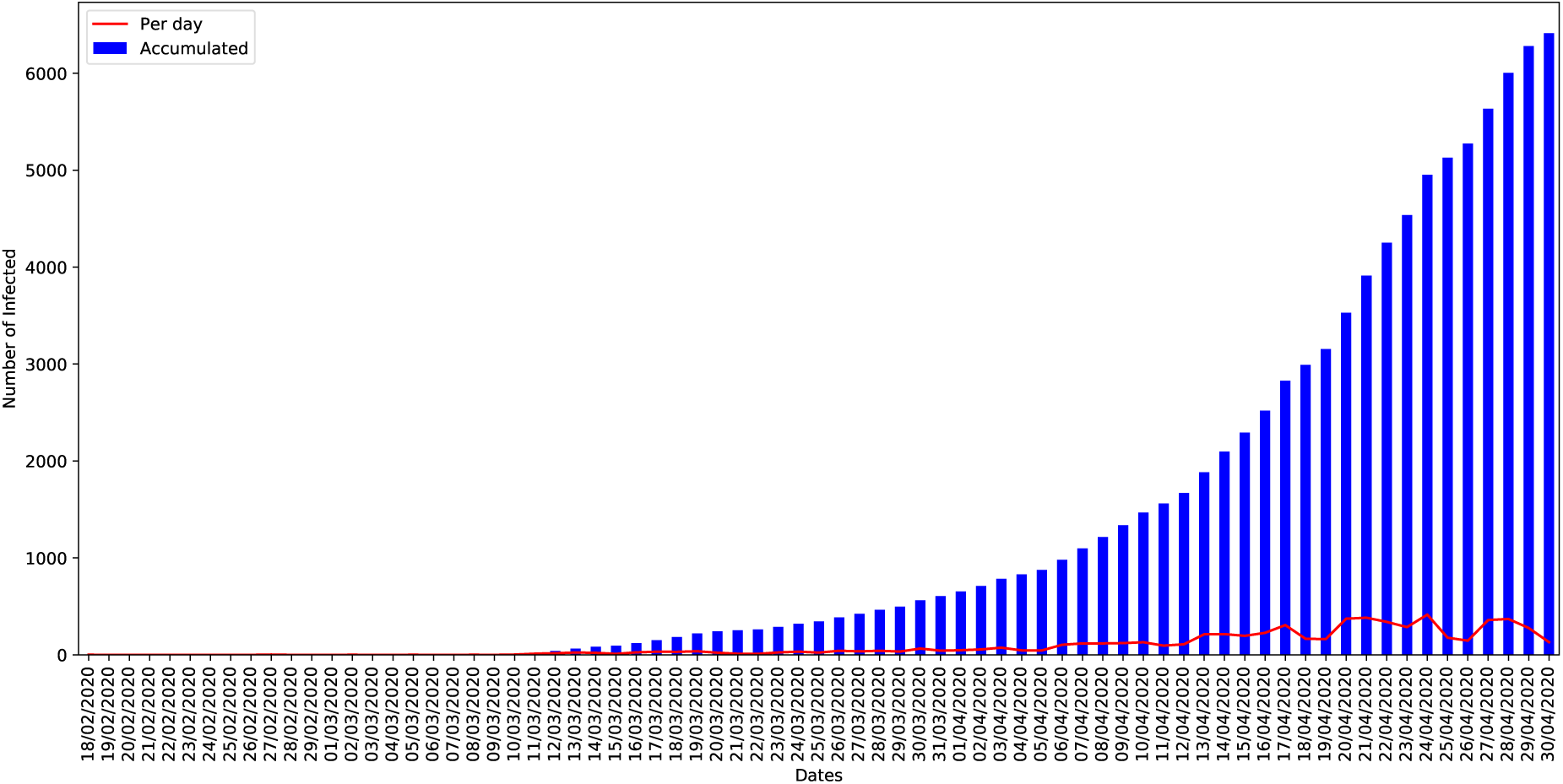
People infected in Mexico City

### B. How often people of Mexico City post about Covid-19 and when did they start?

The tweets or posts during the third week of March from 13-03-2020 to 20-03-2020 were extracted and analyzed in this experiment because in this period of time a significant and constant increase in cases of people infected with Covid-19 were reported. As you can see in the Figure 3, there is a correlation between the increasing number of posts per day and the increasing number of positive cases reported by Mexican government in that time. But, considering the trustability of the post was selected only the top users, and there is a similar distribution than the whole data, the results are presented Figure 4.

**Fig. 3.**
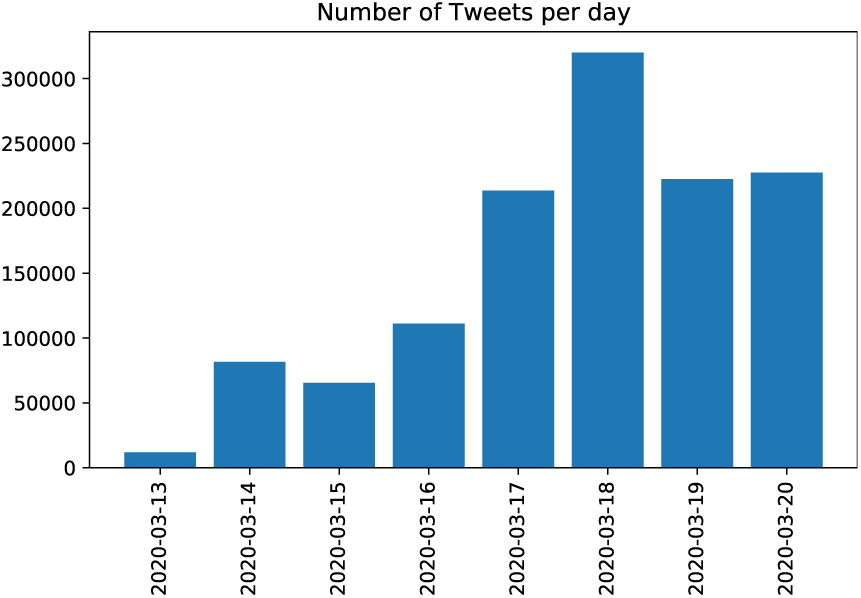
Tweets during the eigth days

**Fig. 4.**
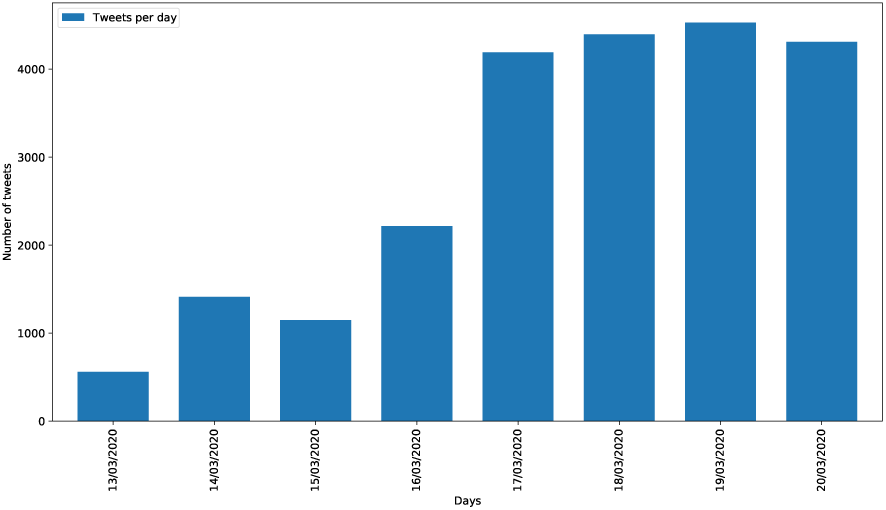
Number of tweets per day in Mexico City.

### C. What is the people of Mexico City posting about Covid-19 on Twitter?

First, Figure 5 shows a cloud of the first thirty terms per day in Mexico City to help identify population concerns related to Covid-19. This experimental scenario is considering all users with post between 13-03-2020 to 20-03-2020.

**Fig. 5.**
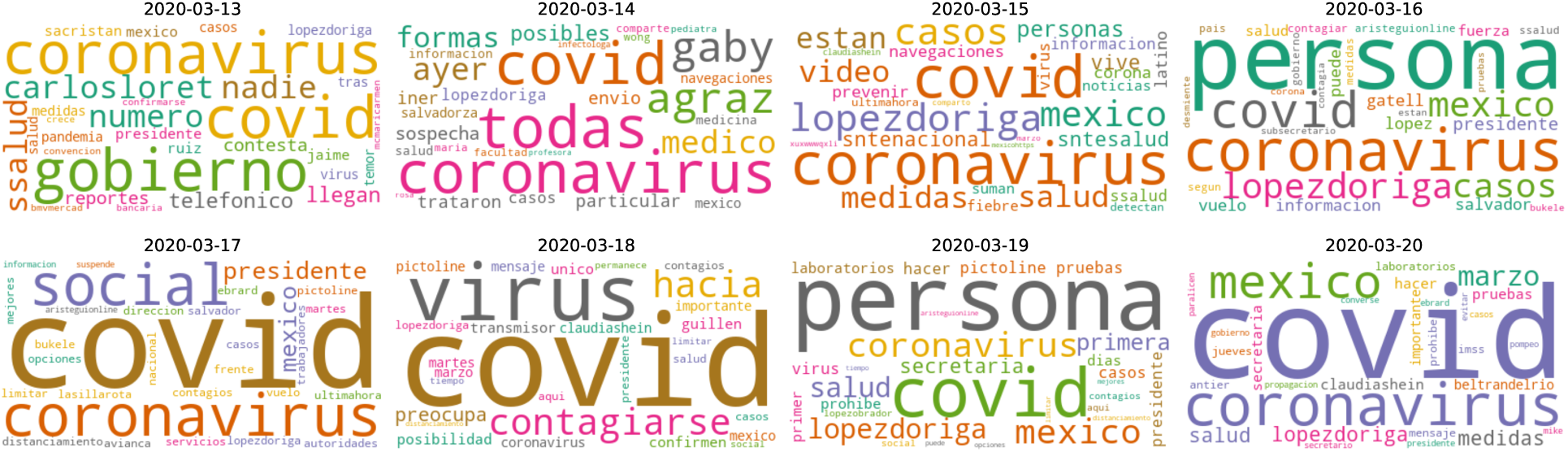
Cloud of words following the histogram of words in Mexico City

Secondly, the Figure 6 displays the fifty of more frequent terms are related to cases of coronavirus in Mexico City. Meanwhile in the Figure 7 shows a cloud of those more frequent terms to help in the visualization and analysis of the extracted information. This experimental scenario is considering only the top users between 13-03-2020 to 20-03-2020.

**Fig. 6.**
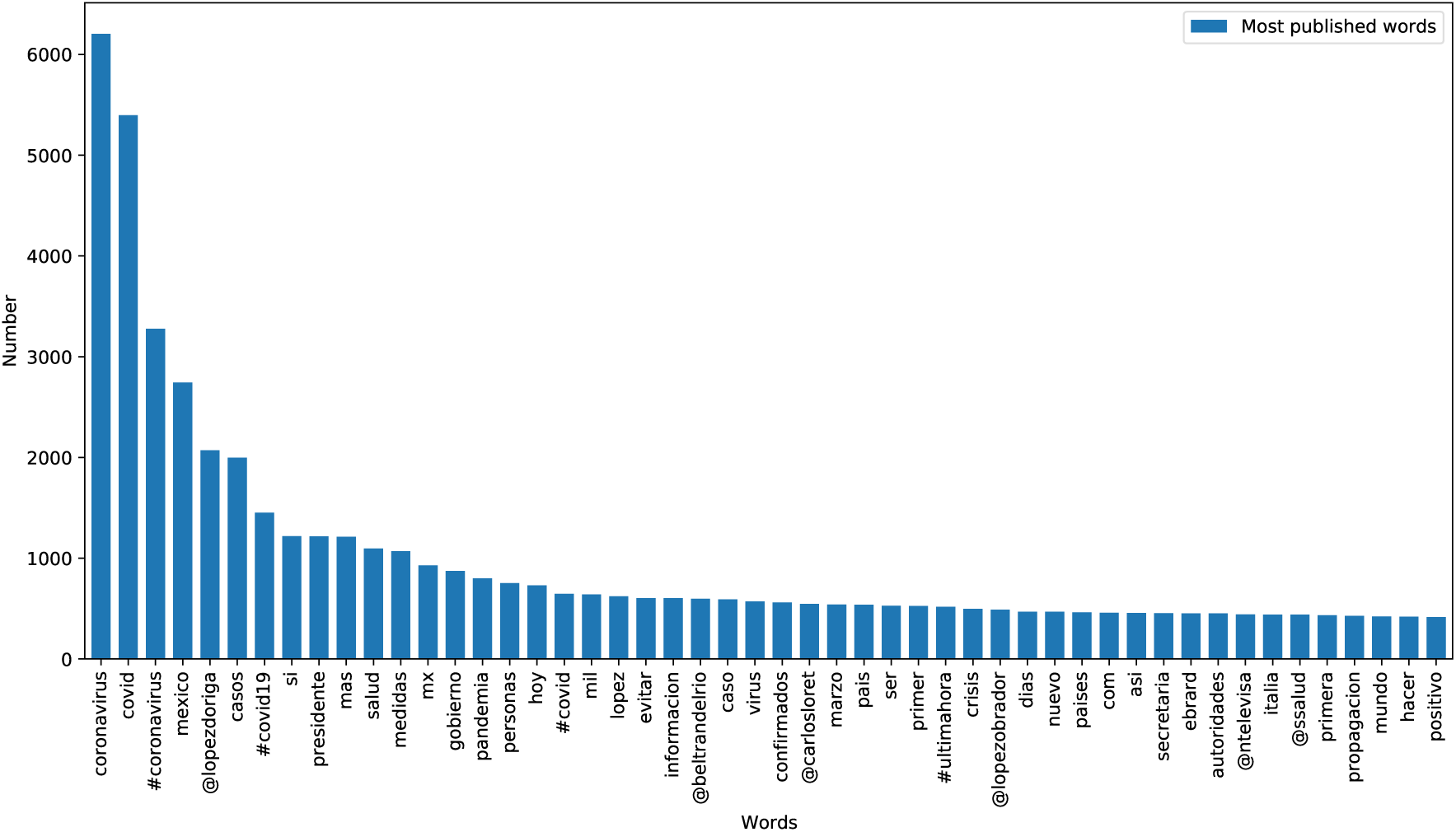
Top 50 of the most posted words

**Fig. 7.**
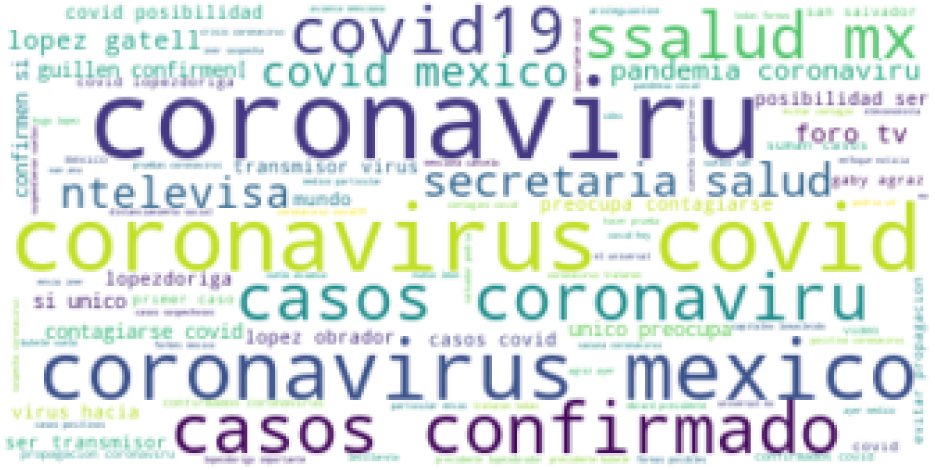
Cloud of Top 50 words in Mexico City on Twitter

After extracting and analyzing a value between number of tweets, in both experimental scenarios of this section, and the frequency of each term, it is concluded that the concerns are related to health risks and economic crisis that could be triggered by Covid-19. It is important to remember that Mexico country promotes a different strategy for the Covid-19 contingency with respect to other countries.

### D. Who are the top 50 users in Mexico City?

It is important and interesting to know who were the most active twitter users during the Covid-19 outbreak in Mexico City. For this experimental scenario only the top users were selected (the same data used in the Figure 4). The Figure 8 shows the names of users and quantity of posts. As can be seen in the graphic, the user accounts with the highest number of tweets are related to the media: written press, radio and television, which they represents 40% the total twitter messages posts between 13-03-2020 and 20-03-2020, with average of 408,6 posts by username in this period.

**Fig. 8.**
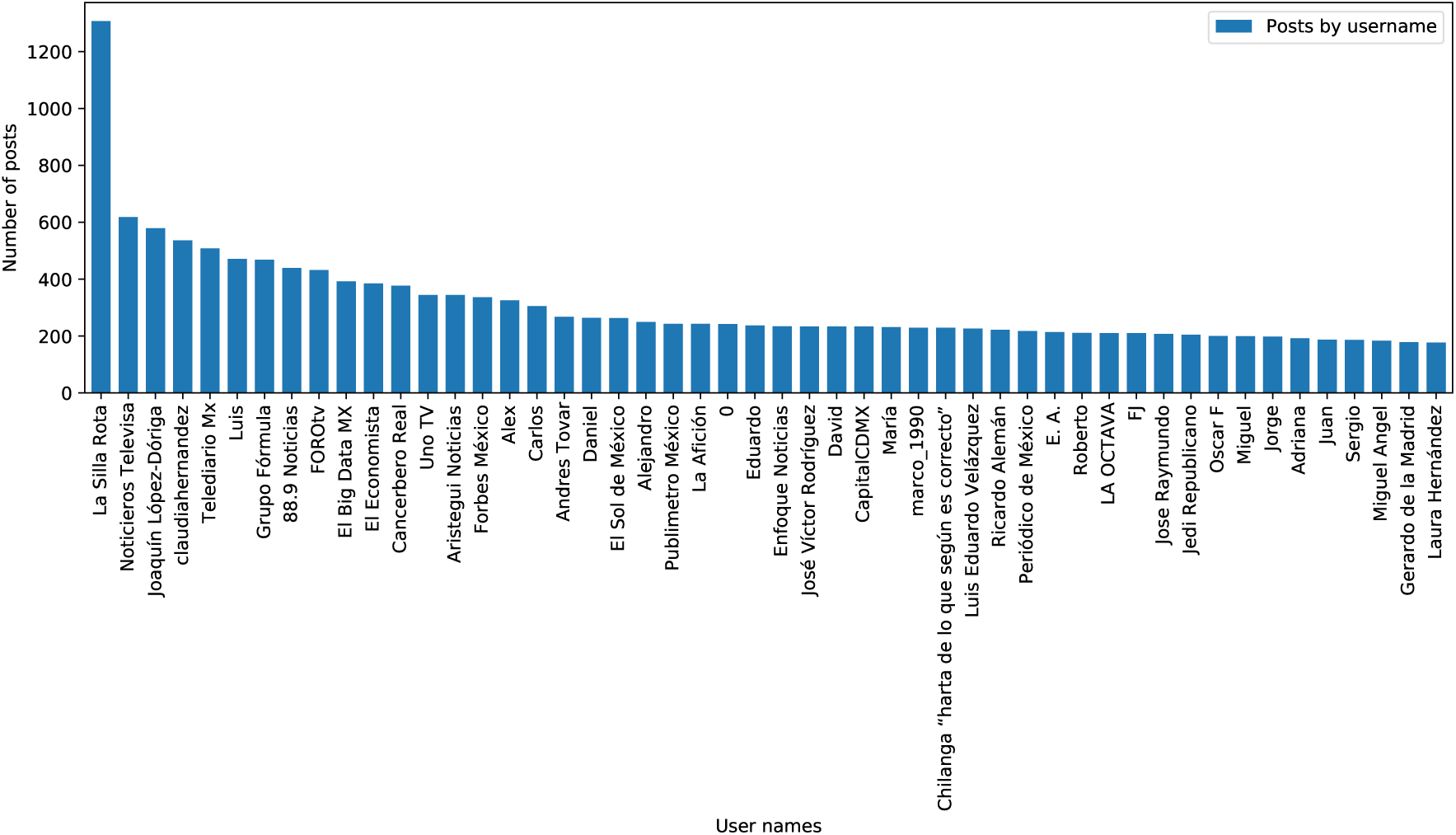
Top 50 of users with most posts in Mexico City

### E. What is the Top 50 users in Mexico City posting about Covid-19 on Twitter?

First, in the Figure 9 is shown the fifty of more frequent terms related to Covid-19 in Mexico City. Meanwhile the Figure 10 shows a cloud of words to help in the visualization and analysis of those more frequent terms. This experimental scenario is considering the Top 50 words published by Top 50 users between 13-03-2020 to 20-03-2020.

**Fig. 9.**
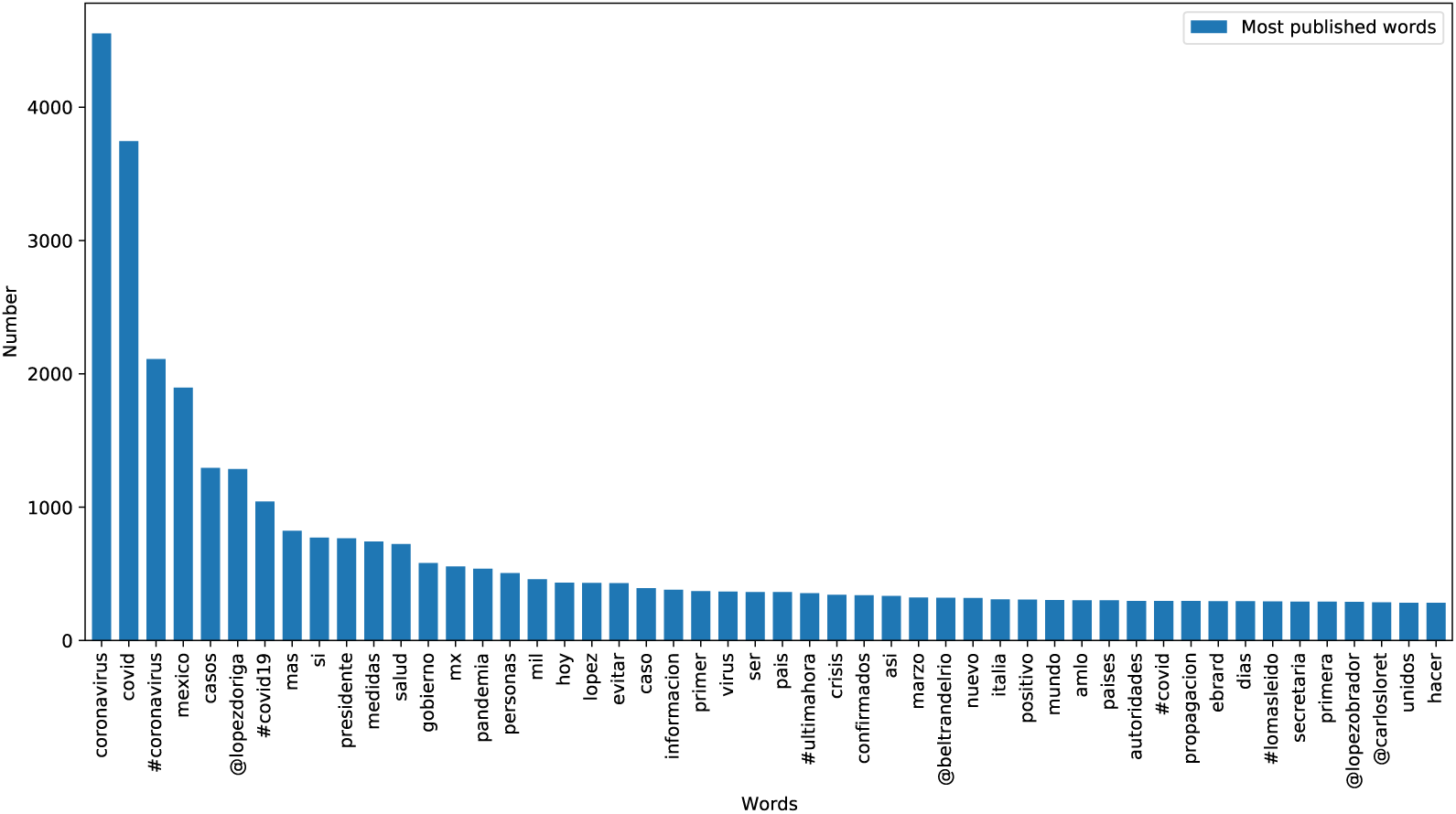
Histogram of Top 50 words published by the Top 50 users in Mexico City

**Fig. 10.**
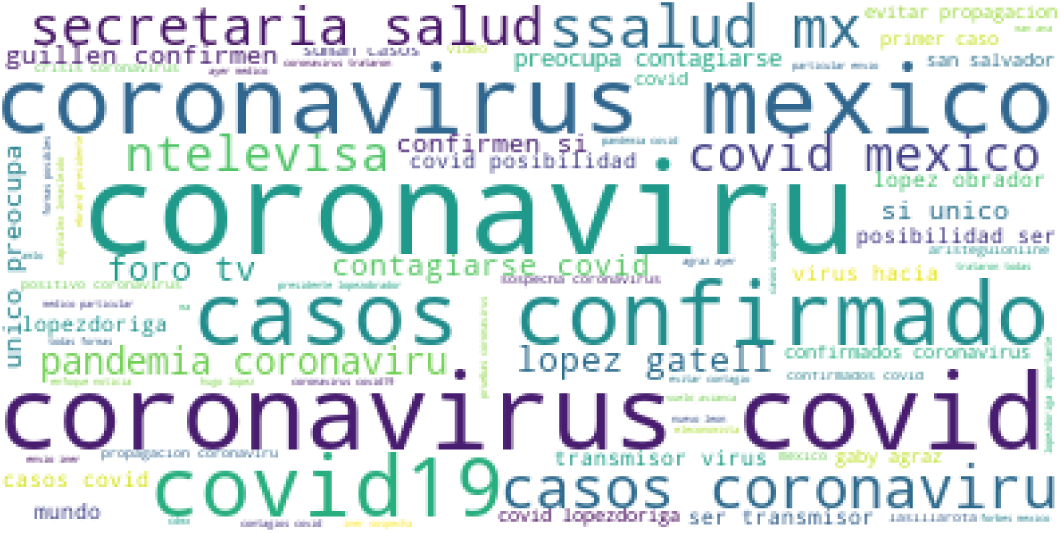
Cloud of words following the histogram of Top 50 words post by Top 50 users in Mexico City on Twitter

Another experimental scenario was to remove media (written press, radio and television) tweets from the Top 50 twitter users in order to know which are the most important topics or concerns related to pandemic and identify if these are same with respect to media, with 6% more posts reported. The experimental results are presented in the Figure 11.

**Fig. 11.**
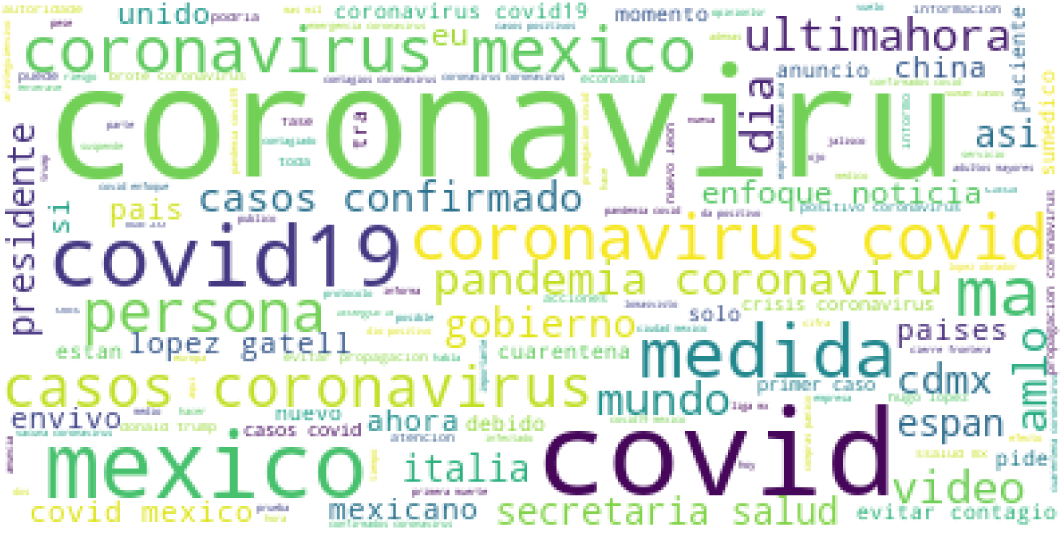
Cloud of words published by the non-media in Mexico City on Twitter

After analyzing experimental results reported in the two previous images it is concluded that the interest topics of population during the outbreaks of Covid19 in Mexico City is about of the first infections in Mexico, health sector strategies to prevent prevention, fear of contagion, among others.

## V. Conclusions

In this paper was presented a Text Mining approach to helps to visualize and understand the impact of Covid-19 in Mexico City population. The experimental scenarios were focused to extract and analyze the most published terms, the most active twitter both from the all data obtained between 13-03-2020 to 20-03-2020 as one proportion of them. It is concluded generally that the central themes of publications related to the information that government authorities officially declared on the number of people infected and strategies designed to mitigate the pandemic, since the latter would affect further the economy the most vulnerable population of Mexico City, one of the most populated cities in the world and Latin America.

## Data Availability

The data will be available soon in a github repository github.com/jecs89

## Notes

### Competing Interest Statement

The authors have declared no competing interest.

### Funding Statement

No funding

